# Use of the FebriDx point-of-care assay as part of a triage algorithm for medical admissions with possible COVID-19

**DOI:** 10.1101/2021.01.05.21249154

**Authors:** Hamish Houston, Gavin Deas, Shivam Naik, Kamal Shah, Shiras Patel, Maria Greca Dottori, Michael Tay, Sarah Filson, James Biggin-Lamming, John Ross, Natalie Vaughan, Nidhi Vaid, Guduru Gopal Rao, Amit K Amin, Ankur Gupta-Wright, Laurence John

## Abstract

**Background:** Patients admitted to hospital with COVID-19 need rapid identification and isolation to prevent nosocomial transmission. However, isolation facilities are often limited, and SARS-CoV-2 RT-PCR results are often not available when discharged from the emergency department. We evaluated a triage algorithm to isolate patients with suspected COVID-19 using simple clinical criteria and the FebriDx assay.

**Design:** Retrospective observational cohort

**Setting:** Large acute care hospital in London, UK

**Participants:** All medical admissions from the ED between 10^th^ August 2020 and 4^th^ November 2020 with valid SARS-CoV-2 RT-PCR.

**Interventions:** Medical admissions were triaged as likely, possible or unlikely COVID-19 based on clinical criteria. Patients triaged as possible COVID-19 underwent FebriDx lateral flow assay on capillary blood, and those positive for MxA were managed as likely COVID-19.

**Primary Outcome measures:** Diagnostic accuracy (sensitivity, specificity and predictive values) of the algorithm and the FebriDx assay compared to SARS-CoV-2 RT-PCR from nasopharyngeal swabs as the reference standard.

**Results:** 4.0% (136/3,443) of medical admissions had RT-PCR confirmed COVID-19. Prevalence of COVID-19 was 45.7% (80/175) in those triaged as likely, 4.1% (50/1,225) in possible and 0.3% (6/2,033) in unlikely COVID-19. Compared to SARS-CoV-2 RT-PCR, clinical triage had sensitivity of 95.6% (95%CI: 90.5% - 98.0%) and specificity of 61.5% (95%CI: 59.8% - 63.1%), whilst the triage algorithm including FebriDx had sensitivity of 92.6% (95%CI: 86.8% - 96.0%) and specificity of 86.4% (95%CI: 85.2% - 87.5%). The triage algorithm reduced the need for 2,859 patients to be admitted to isolation rooms. Ten patients missed by the algorithm had mild or asymptomatic COVID-19.

**Conclusions:** A triage algorithm including FebriDx assay had good sensitivity and was useful to ‘rule-out’ COVID-19 among medical admissions to hospital.

**STRENGTHS AND LIMITATIONS OF THIS STUDY:**

- Pragmatic study including a large cohort of consecutive medical admissions providing routine clinical care.
- A single SARS-CoV-2 RT-PCR is an imperfect reference standard for COVID-19. Multiple RT-PCR platforms used, with different PCR targets and performance.
- A higher prevalence of COVID-19 or other respiratory pathogens might alter performance.
- Criteria for likely and possible COVID-19 groups changed subtly during the study period.

## INTRODUCTION

The Coronavirus disease (COVID-19) pandemic, caused by SARS-CoV-2, presents unprecedented challenges for infection prevention and control (IPC) within healthcare facilities worldwide.^1^ Transmission may occur via respiratory droplet, fomite, or airborne routes (following aerosol-generating procedures).^1–3^ Prolonged indoor contact increases transmission, and nosocomial transmission is common.^4,5^ Respiratory isolation capacity (neutral or negative pressure side-rooms) is easily saturated within healthcare facilities.^6^ Decisions to isolate patients in need of admission with suspected or possible COVID-19 must be rapid and accurate to maintain patient flow from emergency departments (EDs), yet minimise risk of nosocomial transmission.

As COVID-19 can present with non-specific symptoms, diagnostic confirmation is often sought by detection of SARS-CoV-2 ribonucleic acid (RNA) by reverse transcription-polymerase chain reaction (RT-PCR) from nasopharyngeal swab (NPS).^7^ However, decisions about patient isolation from ED are usually required before the results of RT-PCR assays are available.^8,9^ Even near-patient, rapid RT-PCR platforms with assay run times of 1-2 hours can be quickly overwhelmed, especially during peaks of COVID-19 incidence.^10,11^ Multivariable diagnostic risk models, including clinical criteria and thoracic imaging, are not sufficient, but may be useful as a triage test to ration expensive or scarce point-of care assays.^12,13^

FebriDx (Lumos diagnostics, Sarasota, Florida, US) is a lateral flow assay that detects two host response proteins, Myxovirus resistance protein A (MxA, positive if >40ng/mL) and C-reactive protein (CRP, positive if >20mg/L) in capillary blood samples. MxA is an interferon-induced antiviral host response protein that has been studied as a biomarker to differentiate bacterial and viral respiratory infections.^14–17^ More recently FebriDx has demonstrated a sensitivity of 93% and specificity of 86% for detecting COVID-19 compared to RT-PCR.^18^ FebriDx could be useful as an early triage tool to identify patients with COVID-19 and help guide isolation and IPC in patients needing admission to hospital.^18–21^ We therefore developed and implemented a COVID-19 triage algorithm, supported by FebriDx, to inform patient flow from the ED whilst awaiting RT-PCR results. Here we describe the diagnostic performance of this algorithm compared to SARS-CoV-2 RT-PCR. We also describe the impact on isolation room demand and the time to FebriDx and RT-PCR results.

## METHODS

### Patient cohort

We utilised data prospectively entered into a COVID-19 triage database and retrospective extraction of clinical and bed allocation data from electronic patient records and hospital IT systems at Northwick Park Hospital, a large district general hospital serving a diverse population in North-West London. Patients were included if they required admission to a medical ward from the ED between 10^th^ August 2020 and 4^th^ November 2020 inclusive.

Consecutive medical admission were triaged into three categories for their likelihood of COVID-19 (unlikely, possible and likely) according to clinical features, observations and plain chest radiograph by the attending clinician based on Public Health England guidance (Table 1 and Supplementary Table 1).^22^ Patients in the possible group underwent testing with FebriDx unless they declined, were immunosuppressed, required high dependency unit or intensive care unit (HDU/ICU) admission, had symptoms of COVID-19 for more than 10 days or had had COVID-19 previously. All patients underwent NPS testing with SARS-CoV-2 RT-PCR, with rapid RT-PCR assays being prioritised for patients in the likely group.

**Table 1.** Clinical Criteria for determining triage groups, testing strategy and bed allocation from the Emergency Department prior to RT-PCR result. Clinical criteria for determining triage groups are shown as of 08/10/2020. Changes to these criteria over time are detailed in supplementary table 1. * Patients were excluded from FebriDx testing if they had a prior history of COVID-19, were immunosuppressed, required intensive care or high dependency unit admission, or had had COVID-19 symptoms for > 10 days. RT-PCR=Reverse transcription polymerase chain reaction, ED=Emergency department

| COVID-19 triage category | Clinical Criteria | Diagnostics performed in ED | Bed Allocation from ED |
| --- | --- | --- | --- |
| **Likely** | Recent Contact with a confirmed COVID-19 case<br>OR<br>Travel to High Risk country within the last 14 days | Routine RT-PCR | Isolation Room |
|  | Known COVID-19 illness confirmed prior to current attendance | Urgent RT-PCR | COVID-19 cohort area or isolation room |
|  | High Clinical Suspicion (eg. Oxygen Requirement, Bilateral infiltrates, Normal WCC/high CRP)<br>OR<br>Change in Normal sense of Smell or Taste |  | Isolation Room |
| **Possible** | Clinical or Radiological Pneumonia<br>OR<br>Fever / Persistent Cough / Shortness of Breath / Hypoxia / Diarrhoea / Confusion | FebriDx * & Urgent RT-PCR | Isolation Room if FebriDx Positive |
|  |  |  | Non-COVID Area if FebriDx Negative |
| **Unlikely** | None of the Above | Routine RT-PCR | Non-COVID Area |

Patients with confirmed COVID-19 on SARS-CoV-2 RT-PCR, those triaged as likely, and those triaged as possible with a positive FebriDx or unable to have a FebriDx test were admitted to an isolation room or COVID-19 cohort area. Patients assigned to the unlikely COVID-19 group and those with a negative FebriDx test were admitted to ‘non-COVID’ wards whilst awaiting SARS-CoV-2 RT-PCR results. Patients were excluded from the triage system if they were under sixteen years of age or admitted under specialities other than medicine.

FebriDx testing was implemented as part of routine clinical care in response to data on assay performance for COVID-19 and an urgent clinical need.^21^ The study was approved by the London North West University Hospitals Trust Research and Development Committee, and given this was a retrospective review using routinely collected clinical data, they deemed formal ethical approval was not required. Results are reported in compliance with STARD and STROBE guidelines (see supplementary materials).

### Testing procedures and definitions

The FebriDx assay was performed as per the manufacturer’s instructions at the point-of-care by ED health-care assistants following training. In brief, 5µL of capillary blood is placed on the sample window and reagents are released by pressing a button. The result is read after 10 minutes, with a positive result being the presence of a blue line in the control window and a red line in the MxA window (limit of detection 40ng/ml). The results from the CRP window were not used given all patients had laboratory CRP measurements. Staff performing FebriDx had access to clinical information but not SARS CoV-2 RT-PCR results at the time of FebriDx testing. Routine SARS CoV-2 RT-PCR was done on NPS using either the Panther Fusion SARS-CoV-2 (Hologic Inc, CA, USA), Abbott RealTime SARS-CoV-2(Abbott Park, IL, USA) or an extraction-free SARS-CoV-2 RT-PCR assay developed by Health Services Laboratories (HSL), UK.^23^ Rapid RT-PCR assays used were Xpert Xpress SARS-CoV-2 (Cepheid, CA, USA) or SAMBA II SARS-CoV-2 (Diagnostics for the Real World, CA, USA).

Patients were defined as having COVID-19 or not based on the first valid RT-PCR result up to 72 hours after admission. Patients without a valid RT-PCR result or triage status were excluded from the analysis. Vital signs, including National Early Warning Score (NEWS) were recorded on arrival to the ED. All biochemical, haematological and radiological data were from the first results within 48 hours of admission. Thoracic imaging (chest radiographs and CT) were reported and coded based upon guidelines on COVID-19 from the British Society of Thoracic Imaging (BSTI) at the time of reporting by radiologists.^24^ Vital status is reported at the time of hospital discharge or data extraction (20^th^ November 2020) for those who were still inpatients.

### Data Analysis and Statistical Methods

We calculated the proportion of patients with confirmed COVID-19 in each triage category, and the diagnostic accuracy (sensitivity, specificity, and positive and negative predictive values with 95% confidence intervals) of both the triage algorithm overall, and the FebriDx assay in patients with possible COVID-19 compared to a SARS-CoV-2 RT-PCR reference standard. Patients with missing RT-PCR or those missing data on triaging were excluded from analysis. We also reported time to FebriDx testing and valid RT-PCR testing. We described the proportion of patients with COVID-19 who were correctly isolated, estimated the number of isolation beds made available by FebriDx testing, and described the patients with COVID-19 who were incorrectly triaged by the algorithm. Basic descriptive statistics were performed, with comparisons made using chi-squared tests for proportions, t-tests for means and Wilcoxon rank sum for medians. Logistic regression was used to compare age and sex adjusted estimates of in-hospital death in each triage group, using complete cases only. Statistical analyses were performed using Stata version 14.0 (StataCorp, LLC, College Station TX). Based on an anticipated sensitivity of 93%, a sample size of 3335 would estimate the sensitivity of the triage algorithm ±5% with alpha 0.05 and prevalence of 3%.

## RESULTS

### Baseline characteristics and COVID-19 diagnosis

Between the 10^th^ August and 4^th^ November 2020, there were 9,645 emergency department attendances resulting in further hospital care. Of these, 3,433 (35.6%) were adult medical patients admitted for further treatment, were triaged using the algorithm based on COVID-19 status and had a valid SARS-CoV-2 RT-PCR result (figure 1). 175 (5.1%) patients were triaged as likely COVID-19, 2,033 (59.2%) patients as unlikely and 1,225 (35.7%) patients were triaged into the possible COVID-19 category. Key patient characteristics are given in Table 2.

**Table 2:** Baseline characteristics, vital signs, initial investigations, mortality and SARS-CoV-2 RT-PCR results for patients in the unlikely, possible and likely COVID-19 groups. For observations on arrival, 3.2 to 4.1% of data were missing. Data were missing for 5.5% of CRP results and 4.0% of haematology results, 22.4% of chest radiograph reports and 2.1% of discharge outcomes. 96 patients (2.8%) had a chest CT report available. Imaging reports were coded as per BSTI guidelines. Chest Radiograph reports were coded as: CVCX0 = Normal; CVCX1 = Classic; CVCX2 = Indeterminate; CVCX3 = Non-COVID-19. Chest CT reports were coded as: CVCT0= Normal; CVCT1= Classic/probable; CVCT2= Indeterminate; CVCT3= Non-COVID-19. Pair-wise comparisons were performed using chi-squared tests for proportions, t-tests for means and Wilcoxon rank sum for median. *P-values are shown for the comparison between the possible and likely COVID-19 groups IQR=Inter-quartile range, CI=Confidence Interval, NEWS=National Early Warning Score, SpO2=Oxygen Saturations, CRP=C-Reactive Protein, CT=Computerised Tomography

| Variable | Unlikely | Possible | Likely | P-value* |
| --- | --- | --- | --- | --- |
| N | 2033 | 1225 | 175 |  |
| Age (years) median (IQR) | 69 (49, 82) | 75 (60, 84) | 62 (48, 74) | <0.001 |
| Age over 65 years, n (%; 95%CI) | 1128 (55.5%, 53.3; 57.6) | 846 (69.1%, 66.5; 71.6) | 79 (45.1%, 37.8; 52.5) | <0.001 |
| Female Sex, n (%; 95%CI) | 969 (47.7%, 45.5; 49.8) | 603 (49.2%, 46.4; 52.0) | 72 (41.1%, 33.9; 48.4) | 0.045 |
| Male Sex, n (%; 95%CI) | 1064 (52.3%, 50.2; 54.5) | 622 (50.8%, 48.0; 53.6) | 103 (58.9%, 51.6; 66.1) |  |
| NEWS, median (IQR) | 1 (0, 3) | 4 (2, 6) | 5 (3, 7) | 0.017 |
| Respiratory Rate (breaths/min), median (IQR) | 18 (18, 20) | 24 (20, 28) | 24 (21, 32) | <0.001 |
| SpO2 <94%, n (%; 95%CI) | 61 (3.1%, 2.4; 3.9) | 234 (19.5%, 17.3; 21.8) | 38 (22.2%, 16.0; 28.5) | 0.41 |
| Required Supplemental Oxygen, n (%; 95%CI) | 52 (2.7%, 2.0; 3.4) | 245 (20.4%, 18.1; 22.7) | 52 (30.4%, 23.5; 37.3) | 0.003 |
| Temperature >37.5°C, n (%; 95%CI) | 172 (8.8%, 7.6; 10.1) | 359 (30.0%, 27.4; 32.6) | 73 (42.7%, 35.3; 50.1) | <0.001 |
| Chest Radiograph - Normal, n (%; 95%CI) | 1171 (81.0%, 79.0; 83.0) | 537 (49.9%, 46.9; 52.9) | 42 (29.8%, 22.2; 37.3) | <0.001 |
| Chest Radiograph - Typical for COVID-19, n (%; 95%CI) | 4 (0.3%, 0.0; 0.5) | 25 (2.3%, 1.4; 3.2) | 54 (38.3%, 30.3; 46.3) | <0.001 |
| Chest Radiograph - Other, n (%; 95%CI) | 271 (18.7%, 16.7; 20.8) | 514 (47.8%, 44.8; 50.8) | 45 (31.9%, 24.2; 39.6) | <0.001 |
| Chest CT - Normal, n (%; 95%CI) | 8 (23.5%, 9.3; 37.8) | 9 (16.4%, 6.6; 26.1) | 0 (0.0%, 0.0; 0.0) | 0.25 |
| Chest CT - Typical for COVID-19, n (%; 95%CI) | 0 (0.0%, 0.0; 0.0) | 3 (5.4%, -0.5; 11.5) | 3 (42.9%, 6.2; 79.5) | 0.002 |
| Chest CT - Other, n (%; 95%CI) | 26 (76.5%, 62.2; 90.7) | 43 (78.2%, 67.3; 89.1) | 4 (57.1%, 20.5; 93.8) | 0.22 |
| CRP (mg/L), median (IQR) | 5.7 (1.4, 26.9) | 26.4 (7.05, 87.65) | 53.7 (25.9, 122.7) | <0.001 |
| CRP >20mg/L, n (%; 95%CI) | 545 (28.7%, 26.7; 30.7) | 656 (55.8%, 52.9; 58.6) | 134 (80.2%, 74.2; 86.3) | <0.001 |
| Lymphocyte Count <1.0x10<sup>9</sup>/l, n (%; 95%CI) | 373 (25.3%, 23.1; 27.5) | 383 (43.9%, 40.6; 47.2) | 70 (54.7%, 46.1; 63.3) | 0.022 |
| Neutrophil Count >7.5x10<sup>9</sup>/l, n (%; 95%CI) | 620 (32.0%, 29.9; 34.0) | 598 (50.4%, 47.6; 53.3) | 61 (36.1%, 28.9; 43.3) | <0.001 |
| Crude In Hospital Mortality, n (%; 95%CI) | 57 (2.8%, 2.1; 3.6) | 89 (7.5%, 6.0; 9.0) | 13 (7.8%, 3.7; 11.8) | 0.89 |
| SARS-CoV-2 RNA Detectable on RT-PCR, n (%; 95%CI) | 6 (0.3%, 0.1; 0.5) | 50 (4.1%, 3.0; 5.2) | 80 (45.7%, 38.3; 53.1) | <0.001 |

**Figure 1:**
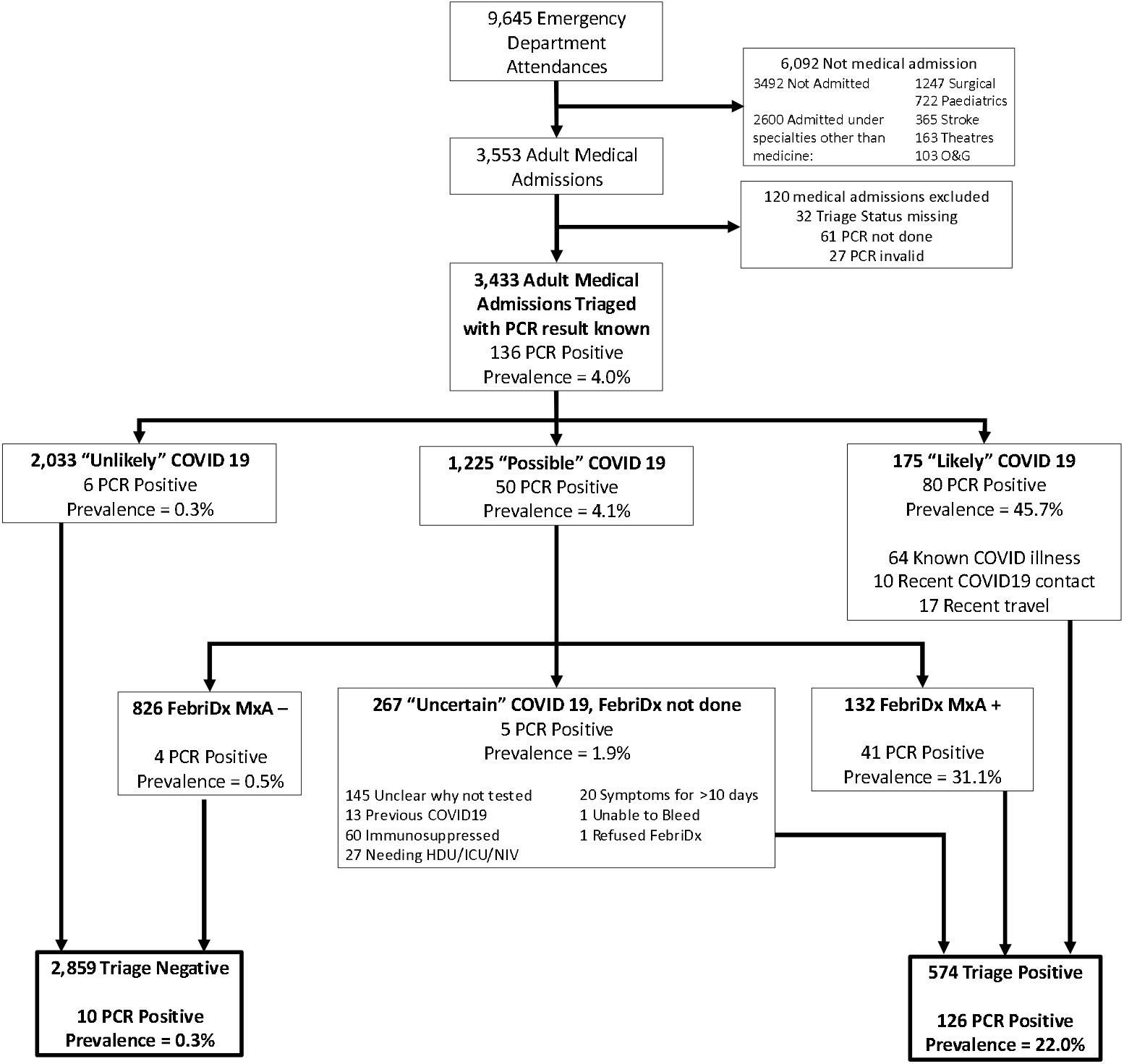
Patient flow through the study and the COVID-19 triage algorithm. Patients were included if they required admission to a medical ward from the ED between 10^th^ August 2020 and 4^th^ November 2020 inclusive. Patients were excluded if they were under sixteen years of age, admitted under specialities other than medicine, or if their triage status or SARS-CoV-2 RT-PCR result was unknown. PCR = SARS-CoV-2 RT-PCR.

There were several differences between the three triage groups (Table 2). The likely COVID-19 group were younger and more unwell at admission (NEWS of 5 vs 1 for patients in the unlikely group, p<0.001) and more frequently required supplemental oxygen (30.4% compared to 2.1% in the unlikely [p<0.001], and 20.4% in the possible group [p=0.003]). As expected, more patients in the likely COVID-19 group had chest radiograph changes typical for COVID-19 than in the other groups (38.3% compared to 2.3% in possible [p<0.001], and 0.3% in unlikely [p<0.001]). The possible COVID-19 group were older (median 75 years [IQR: 60 – 84]) than the other two groups and were more likely to have an elevated neutrophil count (greater than 7.5×10^9/l) than the likely or possible groups.

Overall, 136/3,443 admissions (4.0%) were diagnosed with PCR-confirmed COVID-19. Prevalence of COVID-19 was 45.7% (80/175) in likely patients, and 4.1% (50/1,225) in the possible group. Of those triaged as unlikely COVID-19, only 6/2,033 (0.3%) were SARS-CoV-2 RT-PCR positive.

### Performance of FebriDx and triage algorithm

The overall diagnostic performance of the clinical triage algorithm compared to the gold standard of SARS-CoV-2 RT-PCR is summarised in Table 3. 958 (78.2%) patients in the possible group were tested using FebriDx (figure 1 shows those excluded). 13.8% (132/958) of FebriDx test results were positive for MxA, with 86.2% negative and no invalid results. The median duration of COVID-19 symptoms in patients tested by FebriDx was 2 days (IQR 1-3, n=847). Patients with positive FebriDx results were younger, more likely to be febrile and less likely to have raised neutrophil counts than FebriDx negative patients (supplementary table 2).

**Table 3:** Measures of Diagnostic Performance for the Triage Algorithm (with and without FebriDx) and FebriDx assay alone for the detection of COVID-19, compared to the reference standard of SARS-CoV-2 RT-PCR. Diagnostic performance measures are shown for three tests: the whole triage algorithm including the FebriDx test (with patients in the likely group, those with positive FebriDx results or those in the possible group who were not tested by FebriDx classified as “triage positive” as shown in figure 1.) the whole triage algorithm without FebriDx (with patients in the likely or possible group classified as “triage positive”. CI = Confidence Interval

|  | Algorithm with FebriDx (n = 3433) |  | Algorithm without FebriDx (n=3433) |  | FebriDx only (n = 958) |  |
| --- | --- | --- | --- | --- | --- | --- |
|  | n/N | % (95%CI) | n/N | % (95%CI) | n/N | % (95%CI) |
| Sensitivity | 126 / 136 | 92.6 (86.8 - 96.0) | 130 / 136 | 95.6 (90.5 – 98.0) | 41 / 45 | 91.1 (78.4 - 96.7) |
| Specificity | 2849 / 3297 | 86.4 (85.2 - 87.5) | 2027 / 3297 | 61.5 (59.8 – 63.1) | 822 / 913 | 90.0 (87.9 - 91.8) |
| Negative Predictive Value | 2849 / 2859 | 99.7 (99.4 - 99.8) | 2027 / 2033 | 99.7 (99.3 – 99.9) | 822 / 826 | 99.5 (98.7 - 99.8) |
| Positive Predictive Value | 126 / 574 | 22.0 (18.8 - 25.5) | 130 / 1400 | 9.3 (7.9 – 10.9) | 41 / 132 | 31.1 (23.7 - 39.5) |

31.1% (41/132) of patients with a positive FebriDx had a positive SARS-CoV-2 RT-PCR, whilst only 4/826 (0.5%) with a negative FebriDx were diagnosed as having COVID-19. All 4 patients with false-negative FebriDx results had normal chest radiographs. 2 patients tested negative for COVID-19 by SARS-CoV-2 RT-PCR but had positive FebriDx results and chest radiograph appearances typical for COVID-19. In the possible COVID-19 group, FebriDx results were available a median of 2.2 hours (IQR: 1.4 to 3.1, n=808) and RT-PCR results a median of 17.8 hours (IQR: 11.35 – 25.34, n=456) after arrival to the ED (figure 2). 88.0% of FebriDx results were available within 4 hours of arrival (n=808).

**Figure 2:**
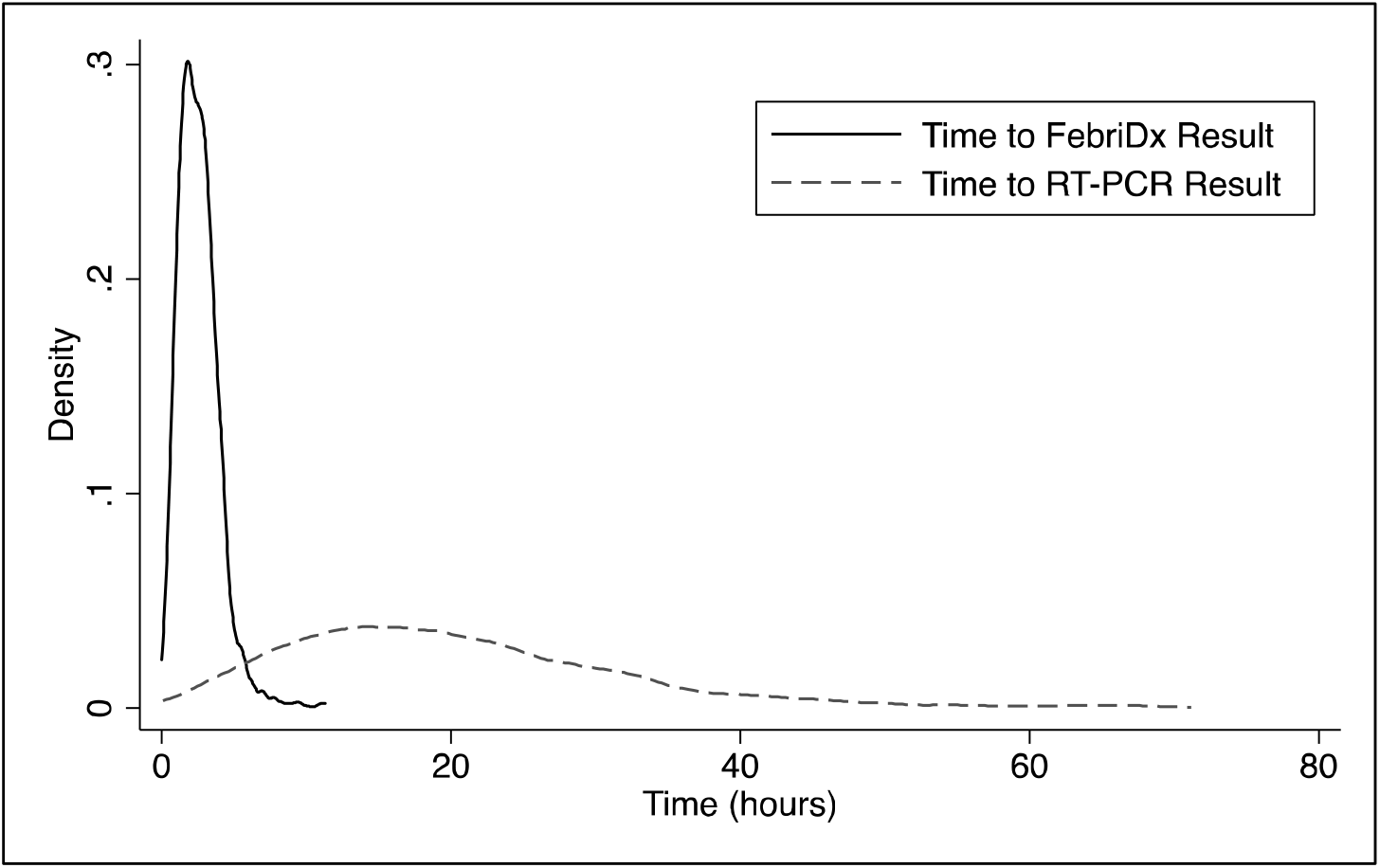
Time from arrival to the availability of FebriDx and SARS-CoV-2 RT-PCR results. Kernel frequency density plot using the Epanechniko function; Time to FebriDx result was calculated as the time from arrival to the emergency department until the time the FebriDx result was recorded (blue plot), bandwidth=0.3; Time to RT-PCR result was calculated as the time from arrival to to the emergency department until the time the SARS-CoV-2 RT-PCR result was recorded (red plot), bandwidth=2.

The triage algorithm correctly identified 126/136 patients with PCR-confirmed COVID-19 in the likely group (sensitivity 92.6%, 95%CI: 86.8 - 96.0) (Table 3 and Supplementary Table 4). The 10 patients who were SARS-CoV-2 RT-PCR positive but missed by the triage algorithm are described in supplementary table 3. 6/10 were classified as unlikely, and 4/10 were classified as possible COVID-19 and had a negative FebriDx. 2/10 were febrile on admission, none required supplemental oxygen, length of stay was short (median 2 days) and 8/8 had normal chest radiographs (2 did not have thoracic imaging done). Specificity of the algorithm was 86.4% (85.2 - 87.5), and negative predictive value was 99.7% (99.4 - 99.8).

### Outcomes

94.9% (129/136) of patients with COVID-19 were appropriately managed in isolation rooms as a result of the triage algorithm (supplementary table 5). Of the 10 patients with PCR-confirmed COVID-19 not identified by the triage algorithm, only 7 were not managed in an isolation room. Had all patients been isolated until SARS-CoV-2 RT-PCR result was available (ie without using any triage algorithm) 2,859 more isolation rooms would have been used. The FebriDx assay allowed 826 more patients to be managed in ‘non-COVID’ areas than if all patients triaged possible COVID-19 had required isolation (9.5 isolation rooms saved per day).

11 (8.1%) patients with COVID-19 died compared to 150 (4.5%) without COVID-19 (p=0.042). Age and sex adjusted odds of death during the admission were higher for patients in the likely (OR: 3.42, 95% CI: 1.81 - 6.45) and possible groups (OR: 2.44, 95% CI: 1.73 - 3.44) than the unlikely COVID-19 group.

## DISCUSSION

Our main findings are that a pragmatic triage algorithm using simple clinical parameters available within the ED and the FebriDx point-of-care test had good sensitivity (92.6%) and excellent NPV (99.7%) for COVID-19 diagnosed by RT-PCR. Inclusion of FebriDx improved the specificity of triage with minimal reductions in sensitivity, allowing a substantial reduction in the number of isolation rooms needed.

Although clinicians were able to identify patients likely and unlikely to have COVID-19 (45.7% and 0.3% of whom had confirmed COVID-19 respectively) based on clinical assessment, radiology and basic blood tests, their assessment was not sufficiently specific. Patients identified as ‘possible’ COVID-19 still had a 4% prevalence of COVID-19, and were a large enough group to overwhelm isolation room capacity. We demonstrate a simple, rapid test performed at the point-of-care can help further risk stratify this group. In real-life settings in a busy ED, a point-of-care test was able to inform isolation decisions within 4 hours of arrival compared to PCR results which were too slow to inform patient flow from ED, even when using ‘rapid’ PCR assays. Although formal cost-effectiveness analysis was not performed, each FebriDx test only costs about US$18, and this may lead to cost savings.

The strengths of this study are its pragmatic design under routine clinical settings, and that we are able to account for over 95% of medical admissions, reducing risks of bias. There are, however, several limitations. A single SARS-CoV-2 RT-PCR is an imperfect reference standard, and does not account for RT-PCR negative COVID-19 patients. We used multiple RT-PCR platforms, which will have different PCR targets and performance. 10% of patients in the possible group did not get tested with FebriDx for unclear reasons, potentially introducing bias. The prevalence of COVID-19 was 4% in this cohort, and it is unclear what impact a higher prevalence of COVID-19 or other respiratory pathogens such as influenza would have on these findings. The criteria for likely and possible COVID-19 groups changed subtly during the study period, although this is unlikely to significantly alter the outcomes.

These data build on previous studies of FebriDx showing good sensitivity, and utility as a ‘rule-out’ test for COVID-19.^17–20^ We may have underestimated the sensitivity by not testing those patients deemed most likely to have COVID-19, although testing this group would have been unlikely to alter clinical decisions, even if FebriDx negative, given the high pre-test probability. The FebriDx test allowed patients with possible COVID-19 to be divided into two groups with similar characteristics and clinical features, but vastly different COVID-19 prevalence (0.5% in FebriDx negative, and 31.1% in FebriDx positive). However, about 10% of patients in this group were not eligible for FebriDx testing.

Only ten patients with COVID-19 were incorrectly triaged by the algorithm, four of whom were tested and ‘missed’ using FebriDx. These patients were younger, less symptomatic, did not have chest radiograph changes, and mostly likely had mild or asymptomatic COVID-19 infection. Given that MxA is an intracellular GTPase induced by type I and type III interferon responses, it is plausible that sensitivity would be lower in pauci- or asymptomatic infection.^25^ Although the patients missed by the algorithm are potential sources of nosocomial transmission, asymptomatic disease is thought to be less transmissible.^26^ We found no nosocomial cases related to these patients.

In conclusion, we demonstrate a simple triage system including the novel FebriDx point-of-care test had good sensitivity and negative predictive value for COVID-19 and utility for managing medical admissions from the ED.

## Data Availability

Data are available upon reasonable request, subject to approval by the London North West University Healthcare NHS Trust Research and Governance Department and approval from relevant ethics and regulatory bodies.

## Acknowledgements

We would like to acknowledge to all the clinical staff at Northwick Park Hospital who cared for the patients involved in this study. In particular, we give thanks to the point-of-care team for their outstanding work in establishing and running the new point-of-care testing service in the emergency department.

## Author contributions

HH, AGW, LJ, SF, JBL, JR, N Vaughan, N Vaid, GGR, and AKA made substantial contribution to the conception of the work. HH, AGW, LJ, and GD made substantial contribution to the design of the work. HH, GD, SN, KS, SP, MGD, and MT contributed to data acquisition. HH and AGW analysed the data. HH, AGW and LJ contributed to data interpretation. HH and AGW drafted the manuscript. All authors contributed to revising the manuscript critically for important intellectual content, approved the final manuscript and are accountable for all aspects of the work.

## Patient and Public Involvement Statement

Due to the retrospective nature of this study, undertaken during the COVID-19 pandemic, patients or the public were not involved in the design, or conduct, or reporting, or dissemination plans of our research.

## References

1 Hui KPY, Cheung MC, Perera RAPM, et al. Tropism, replication competence, and innate immune responses of the coronavirus SARS-CoV-2 in human respiratory tract and conjunctiva: an analysis in ex-vivo and in-vitro cultures. Lancet Respir Med 2020;8:687–95. doi:10.1016/S2213-2600(20)30193-4

2 Zhou J, Otter JA, Price JR, et al. Investigating SARS-CoV-2 surface and air contamination in an acute healthcare setting during the peak of the COVID-19 pandemic in London. Clin Infect Dis Published Online First: 8 July 2020. doi:10.1093/cid/ciaa905

3 van Doremalen N, Bushmaker T, Morris DH, et al. Aerosol and Surface Stability of SARS-CoV-2 as Compared with SARS-CoV-1. N Engl J Med 2020;382:1564–7. doi:10.1056/nejmc2004973

4 Cevik M, Marcus JL, Buckee C, et al. Severe Acute Respiratory Syndrome Coronavirus 2 (SARS-CoV-2) Transmission Dynamics Should Inform Policy. Clin Infect Dis Published Online First: 23 September 2020. doi:10.1093/cid/ciaa1442

5 Rickman HM, Rampling T, Shaw K, et al. Nosocomial Transmission of Coronavirus Disease 2019: A Retrospective Study of 66 Hospital-acquired Cases in a London Teaching Hospital. Clin Infect Dis Published Online First: 20 June 2020. doi:10.1093/cid/ciaa816

6 Verelst F, Kuylen E, Beutels P. Indications for healthcare surge capacity in European countries facing an exponential increase in coronavirus disease (COVID-19) cases, March 2020. Eurosurveillance 2020;25:1. doi:10.2807/1560-7917.ES.2020.25.13.2000323

7 Wang D, Hu B, Hu C, et al. Clinical Characteristics of 138 Hospitalized Patients with 2019 Novel Coronavirus-Infected Pneumonia in Wuhan, China. JAMA -J Am Med Assoc 2020;323:1061–9. doi:10.1001/jama.2020.1585

8 Brendish NJ, Poole S, Naidu V V., et al. Clinical impact of molecular point-of-care testing for suspected COVID-19 in hospital (COV-19POC): a prospective, interventional, non-randomised, controlled study. Lancet Respir Med 2020;8:1192– 200. doi:10.1016/S2213-2600(20)30454-9

9 NHS England. Pillar 1 NHS labs COVID-19 testing turnaround time data: 15 July 2020. 2020. https://www.england.nhs.uk/coronavirus/publication/nhs-labs-covid-19-esting-turnaround-time-data/

10 Gibani MM, Toumazou C, Sohbati M, et al. Assessing a novel, lab-free, point-of-care test for SARS-CoV-2 (CovidNudge): a diagnostic accuracy study. The Lancet Microbe 2020;1:e300–7. doi:10.1016/s2666-5247(20)30121-x

11 Collier DA, Assennato SM, Warne B, et al. Point of Care Nucleic Acid Testing for SARS-CoV-2 in Hospitalized Patients: A Clinical Validation Trial and Implementation Study. Cell Reports Med 2020;1. doi:10.1016/j.xcrm.2020.100062

12 Fink DL, Khan PY, Goldman N, et al. Development and internal validation of a diagnostic prediction model for COVID-19 at time of admission to hospital. QJM An Int J Med Published Online First: 9 November 2020. doi:10.1093/qjmed/hcaa305

13 Wynants L, Van Calster B, Collins GS, et al. Prediction models for diagnosis and prognosis of covid-19: Systematic review and critical appraisal. BMJ 2020;369:18. doi:10.1136/bmj.m1328

14 Engelmann I, Dubos F, Lobert PE, et al. Diagnosis of viral infections using myxovirus resistance protein a (MxA). Pediatrics 2015;135:e985–93. doi:10.1542/peds.2014-1946

15 Nakabayashi M, Adachi Y, Itazawa T, et al. MxA-based recognition of viral illness in febrile children by a whole blood assay. Pediatr Res 2006;60:770–4. doi:10.1203/01.pdr.0000246098.65888.5b

16 Sambursky R, Shapiro N. Evaluation of a combined MxA and CRP point-of-care immunoassay to identify viral and/or bacterial immune response in patients with acute febrile respiratory infection. Eur Clin Respir J 2015;2:28245. doi:10.3402/ecrj.v2.28245

17 Thomas J, Pociute A, Kevalas R, et al. Blood biomarkers differentiating viral versus bacterial pneumonia aetiology: A literature review. Ital. J. Pediatr. 2020;46:4. doi:10.1186/s13052-020-0770-3

18 Clark TW, Brendish NJ, Poole S, et al. Diagnostic accuracy of the FebriDx host response point-of-care test in patients hospitalised with suspected COVID-19. J Infect 2020;81:607–13. doi:10.1016/j.jinf.2020.06.051

19 Karim N, Ashraf MZ, Naeem M, et al. Utility of the FebriDx point-of-care test for rapid triage and identification of possible coronavirus disease 2019 (COVID-19). Int J Clin Pract Published Online First: 17 September 2020. doi:10.1111/ijcp.13702

20 National Institute for Health and Care Excellence. Overview | FebriDx for C-reactive protein and myxovirus resistance protein A testing | Advice | NICE. https://www.nice.org.uk/advice/mib224/ (xaccessed 19 Dec 2020).

21 Ismail SA, Huntley C, Post N, et al. Horses for courses? Assessing the potential value of a surrogate, point-of-care test for SARS-CoV-2 epidemic control. Influenza Other Respi Viruses 2020;00:1– 4. doi:10.1111/irv.12796

22 Public Health England. COVID-19: infection prevention and control guidance. https://www.publichealth.hscni.net/sites/default/files/2020-10/COVID-19_Infection_prevention_and_control_guidance_complete.3.2%2818_06_2020%29_0.pdf

23 Grant P, Turner M, Shin GY, et al. Extraction-free COVID-19 (SARS-CoV-2) diagnosis by RT-PCR to increase capacity for national testing programmes during a pandemic. bioRxiv [Preprint] Published Online First: 9 April 2020. doi:10.1101/2020.04.06.028316

24 British Society of Thoracic Imaging. BSTI COVID-19 CXR Report Proforma. https://www.google.com/url?sa=t&rct=j&q=&esrc=s&source=web&cd=&ved=2ahUKEwj_j4CurtrtAhUnQUEAHRTWA80QFjABegQIARAC&url=https%3A%2F%2Fwww.bsti.org.uk%2Fmedia%2Fresources%2Ffiles%2FBSTI_COVID_CXR_Proforma_v.3-1.pdf&usg=AOvVaw0vUxoui2fz68LEbrs6eTQm

25 Shirley M. FebriDx®: A Rapid Diagnostic Test for Differentiating Bacterial and Viral Aetiologies in Acute Respiratory Infections. Mol Diagnosis Ther 2019;23:803–9. doi:10.1007/s40291-019-00433-x

26 Cevik M, Tate M, Lloyd O, et al. SARS-CoV-2, SARS-CoV, and MERS-CoV viral load dynamics, duration of viral shedding, and infectiousness: a systematic review and meta-analysis. The Lancet Microbe 2020;0. doi:10.1016/s2666-5247(20)30172-5

