## Supplementary Tables for "Use of the FebriDx point-of-care assay as part of a triage algorithm for medical admissions with possible COVID-19": Supplementary Tables 13.1.21.docx

**Supplementary Table 1: Changes made to the inclusion criteria for the triage categories and the exclusion criteria for FebriDx testing during the study period.**

|  | Date of Update to COVID-19 Triage Criteria | | | |
| --- | --- | --- | --- | --- |
|  | 06/07/2020 | 09/09/2020 | 21/09/2020 | 08/10/2020 |
| Likely | Confirmed COVID-19 during current illness | | | |
|  | High Clinical Suspicion  (eg. Oxygen Requirement, Bilateral infiltrates, Normal WCC/high CRP) | | | |
|  | Recent Contact with a confirmed COVID-19 case | | | |
|  | Travel to High Risk country within the last 14 days | | | |
|  |  |  |  | Change in Normal sense of Smell or Taste |
| Possible | Change in Normal sense of Smell or Taste | | | removed |
|  | Clinical or Radiological Pneumonia | | | |
|  | Fever PLUS Persistent Cough OR Shortness of Breath OR Hypoxia | | Fever OR Persistent Cough OR Shortness of Breath OR Hypoxia | |
|  |  |  |  | Confusion OR Diarrhoea |
| Unlikely | None of the Above | | | |
| Exclusion Criteria for FebriDx | Immunosuppressed | | | |
|  | Previous COVID-19 | | | |
|  |  | Requiring ITU/HDU/NIV | | |
|  |  |  | COVID-19 Symptoms >10 days | |

Supplementary Table 1 footnotes:

The inclusion criteria for the triage categories and the exclusion criteria for FebriDx testing were adjusted during the course of this pragmatic study. WCC=white cell count, CRP= C-Reactive Protein, ITU=Intensive Therapy Unit, HDU=High Dependency Unit, NIV=Non-Invasive ventilation.

**Supplementary Table 2: Baseline characteristics, vital signs, initial investigations, mortality and SARS-CoV-2 RT-PCR results for patients in the possible COVID-19 group by FebriDx test result**

| Variable | FebriDx Negative | FebriDx Positive | FebriDx Not Done | FebriDx Positive vs Negative |
| --- | --- | --- | --- | --- |
| N | 826 | 132 | 267 |  |
| Age (years) median (IQR) | 77 (61, 85) | 69.5 (54.5, 81.5) | 72 (60, 81) | <0.001 |
| Age over 65 years, n (%, 95%CI) | 586 (70.9, 67.8; 74.0) | 80 (60.6, 52.0; 68.6) | 180 (67.4, 61.5; 72.8) | 0.017 |
| Female Sex, n (%, 95%CI) | 399 (48.3, 44.9; 51.7) | 63 (47.7, 39.3; 56.3) | 141 (52.8, 46.8; 58.8) | 0.90 |
| Male Sex, n (%, 95%CI) | 427 (51.7, 48.3; 55.1) | 69 (52.3, 43.7; 60.7) | 126 (47.2, 41.2; 53.2) |  |
| NEWS, median (IQR) | 4 (2, 7) | 4 (3, 6) | 4 (2, 6) | 0.62 |
| Respiratory Rate (breaths/min), median (IQR) | 24 (20, 28) | 24 (20, 28) | 24 (19, 28) | 0.74 |
| SpO2 <94%, n (%, 95%CI) | 164 (20.2, 17.6; 23.1) | 24 (18.6, 12.8; 26.3) | 46 (17.9, 13.7; 23.1) | 0.67 |
| Required Supplemental Oxygen, n (%, 95%CI) | 170 (20.9, 18.2; 23.8) | 21 (16.3, 10.8; 23.7) | 54 (21.0, 16.4; 26.5) | 0.23 |
| Temperature >37.5ºC, n (%, 95%CI) | 245 (30.2, 27.1; 33.4) | 55 (42.6, 34.4; 51.4) | 59 (23.0, 18.3; 28.6) | 0.005 |
| Chest Radiograph - Normal, n (%, 95%CI) | 375 (51.2, 47.6; 54.9) | 52 (43.3, 34.7; 52.4) | 110 (49.1, 42.6; 55.7) | <0.001 |
| Chest Radiograph - Typical for COVID-19, n (%, 95%CI) | 8 (1.1, 0.6; 2.2) | 11 (9.2, 5.1; 15.8) | 6 (2.7, 1.2; 5.9) | <0.001 |
| Chest Radiograph - Other, n (%, 95%CI) | 349 (47.7, 44.1; 51.3) | 57 (47.5, 38.7; 56.5) | 108 (48.2, 41.7; 54.8) | 0.97 |
| Chest CT - Normal, n (%, 95%CI) | 9 (19.6, 10.2; 34.1) | 0 (0) | 0 (0) | 0.40 |
| Chest CT - Typical for COVID-19, n (%, 95%CI) | 2 (4.4, 1.0; 16.5) | 0 (0) | 1 (16.7, 0.9; 81.4) | 0.71 |
| Chest CT - Other, n (%, 95%CI) | 35 (76.1, 61.1; 86.5) | 3 (100) | 5 (83.3, 18.6; 99.1) | 0.34 |
| CRP (mg/L), median (IQR) | 26.3 (6.5, 95.5) | 37.05 (17.1, 78.9) | 18.85 (4.9, 76.3) | 0.012 |
| CRP >20mg/L, n (%, 95%CI) | 443 (55.9, 52.5; 59.4) | 87 (66.9, 58.4; 74.5) | 126 (49.6, 43.5; 55.8) | 0.019 |
| Lymphocyte Count <1.0x10^9/l, n (%, 95%CI) | 263 (43.8, 39.9; 47.8) | 46 (47.9, 38.1; 57.9) | 74 (41.8, 34.7; 49.2) | 0.455 |
| Neutrophil Count >7.5x10^9/l, n (%, 95%CI) | 422 (52.8, 49.3; 56.3) | 50 (38.5, 30.5; 47.1) | 126 (49.0, 42.9; 55.2) | 0.002 |
| Mortality, n (%, 95%CI) | 65 (8.1, 6.4; 10.2) | 6 (4.7, 2.1; 10.2) | 18 (6.9, 4.4; 10.8) | 0.19 |
| SARS-CoV2 RNA Detectable on RT-PCR, n (%, 95%CI) | 4 (0.5, 0.3; 1.3) | 41 (31.1, 23.7; 39.5) | 5 (1.9, 0.8; 4.4) | <0.001 |

Supplementary Table 2 footnotes: Missing data are summarised in the footnotes to table 2 in the main text. Imaging reports were coded as per BSTI guidelines. Chest Radiograph reports were coded as: CVCX0 = Normal; CVCX1 = Classic; CVCX2 = Indeterminate; CVCX3 = Non-COVID-19. Chest CT reports were coded as: CVCT0= Normal; CVCT1= Classic/probable; CVCT2= Indeterminate; CVCT3= Non-COVID-19. IQR=Inter-quartile range, CI=Confidence Interval, NEWS=National Early Warning Score, SpO2=Oxygen Saturations, CRP=C-Reactive Protein, CT=Computerised Tomography

**Supplementary Table 3: Baseline characteristics of patients with positive SARS-CoV-2 RT-PCR results who were classified as triage negative by the algorithm**

| Case | 1 | 2 | 3 | 4 | 5 | 6 | 7 | 8 | 9 | 10 |
| --- | --- | --- | --- | --- | --- | --- | --- | --- | --- | --- |
| Triage Status | Unlikely | | | | | | FebriDx Negative | | | |
| Decade of Life* | 5 | 7 | 3 | 5 | 7 | 6 | 6 | 3 | 7 | 5 |
| Sex (F/M) | F | M | F | M | M | M | F | M | F | F |
| Presentation | Fever and epigastric pain | Hypoglycaemic collapse | Hyperkalaemia on clinic bloods | Herpes Zoster | Intentional Overdose | Unstable Angina | URTI symptoms | Diarrhoea | Fever and SOB | Headache and anosmia |
| Duration of Symptoms (days) | x | x | x | x | x | x | 5 | 7 | 1 | 2 |
| NEWS on Arrival | 4 | 1 | 0 | 1 | 2 | 1 | 7 | 2 | 3 | 3 |
| Respiratory Rate (breaths/min) | 20 | 18 | 18 | 20 | 14 | 18 | 32 | 18 | 22 | 21 |
| SpO2 (%) | 97 | 96 | 100 | 96 | 93 | 98 | 94 | 100 | 96 | 100 |
| Required Supplemental Oxygen (Y/N) | N | N | N | N | N | N | N | N | N | N |
| Temperature >37.5ºC, n (%, 95%CI) | 38.1 | 35.2 | 36.5 | 38 | 36.9 | 37 | 39.7 | 38.3 | 38.1 | 36.3 |
| Chest Radiograph | CVCX0 | CVCX0 | ND | CVCX0 | CVCX0 | CVCX0 | CVCX0 | ND | CVCX0 | CVCX0 |
| CRP (mg/L) | 9.5 | 2.6 | 2.6 | 4 | 0.7 | 57.1 | 16.4 | 0.9 | 5.1 | 68.5 |
| Lymphocyte Count (x10^9/l) | 0.5 | 1.4 | 2.2 | 1.1 | 3 | 0.7 | 2.2 | 1.2 | 0.5 | 0.7 |
| Neutrophil Count (x10^9/l) | 8.8 | 9.5 | 6.5 | 2.9 | 2.5 | 1.9 | 6.7 | 2.7 | 4.6 | 1.6 |
| Isolated (Y / N) | N | N | N | Y | N | N | N | Y | Y | N |
| ICU Admission (Y / N) | N | N | N | N | N | N | N | N | N | N |
| Died (Y / N) | N | N | N | N | N | N | N | N | N | N |
| Length of stay (days) | 2 | 1 | 1 | 1 | 7 | 2 | 4 | 2 | 4 | 1 |

Supplementary Table 3 footnotes: *Age on arrival is presented in terms of Decade of Life (eg. 5 = age 40 to 49 years). Imaging reports were coded as per BSTI guidelines. Chest Radiograph reports were coded as: CVCX0 = Normal; CVCX1 = Classic; CVCX2 = Indeterminate; CVCX3 = Non-COVID-19, NEWS=National Early Warning Score, SpO2=Oxygen Saturations, CRP=C-Reactive Protein, Y=Yes, N=No, ND=Not Done

**Supplementary Table 4:**

|  |  | SARS-CoV-2 RT-PCR | | |
| --- | --- | --- | --- | --- |
| A |  | Positive | Negative | Total |
| Algorithm with FebriDx (n=3433) | Positive | 126 | 448 | 574 |
|  | Negative | 10 | 2849 | 2859 |
|  | Total | 136 | 3297 | 3433 |
|  |  | SARS-CoV-2 RT-PCR | | |
| B |  | Positive | Negative | Total |
| Algorithm without FebriDx (n=3433) | Positive | 130 | 1270 | 1400 |
|  | Negative | 6 | 2027 | 2033 |
|  | Total | 136 | 3297 | 3433 |
|  |  | SARS-CoV-2 RT-PCR | | |
| C |  | Positive | Negative | Total |
| FebriDx only (n=958) | Positive | 41 | 4 | 45 |
|  | Negative | 4 | 822 | 826 |
|  | Total | 45 | 913 | 958 |

Table 4 footnotes: Cross tabulation of results of the triage algorithm with FebriDx (A) and without FebriDx (B) as well as the results of FebriDx within the possible COVID-19 group receiving a FebriDx test (C) compared to a SARS-CoV-2 RT-PCR reference standard.

**Supplementary Table 5: Number of patients allocated to isolation rooms or COVID-19 cohorts in SARS-CoV-2 RT-PCR positive patients, and those requiring isolation following triage.**

|  | SARS-CoV-2 RT-PCR Positive (n=136) | Triage Positive (n=574) | Likely  (n=175) | Possible, FebriDx Positive  (n=132) | Possible, FebriDx Not Done  (n=267) |
| --- | --- | --- | --- | --- | --- |
| ‘Non-COVID’ Area | 7 | 68 | 5 | 4 | 58 |
| Side Room | 112 | 477 | 152 | 122 | 203 |
| COVID-19 Cohort Ward | 17 | 29 | 18 | 6 | 6 |
| % Isolated | 94.9 | 88.2 | 97.1 | 97.0 | 78.3 |

Table 5 footnotes: Actual patient movement from the emergency department extracted from the hospital’s bed management system.

STARD 2015

AIM

STARD stands for “Standards for Reporting Diagnostic accuracy studies”. This list of items was developed to contribute to the completeness and transparency of reporting of diagnostic accuracy studies. Authors can use the list to write informative study reports. Editors and peer-reviewers can use it to evaluate whether the information has been included in manuscripts submitted for publication.

Explanation

A **diagnostic accuracy study** evaluates the ability of one or more medical tests to correctly classify study participants as having a **target condition.** This can be a disease, a disease stage, response or benefit from therapy, or an event or condition in the future. A medical test can be an imaging procedure, a laboratory test, elements from history and physical examination, a combination of these, or any other method for collecting information about the current health status of a patient.

The test whose accuracy is evaluated is called **index test.** A study can evaluate the accuracy of one or more index tests. Evaluating the ability of a medical test to correctly classify patients is typically done by comparing the distribution of the index test results with those of the **reference standard**. The reference standard is the best available method for establishing the presence or absence of the target condition. An accuracy study can rely on one or more reference standards.

If test results are categorized as either positive or negative, the cross tabulation of the index test results against those of the reference standard can be used to estimate the **sensitivity** of the index test (the proportion of participants *with* the target condition who have a positive index test), and its **specificity** (the proportion *without* the target condition who have a negative index test). From this cross tabulation (sometimes referred to as the contingency or “2x2” table), several other accuracy statistics can be estimated, such as the positive and negative **predictive values** of the test. Confidence intervals around estimates of accuracy can then be calculated to quantify the statistical **precision** of the measurements.

If the index test results can take more than two values, categorization of test results as positive or negative requires a **test positivity cut-off**. When multiple such cut-offs can be defined, authors can report a receiver operating characteristic (ROC) curve which graphically represents the combination of sensitivity and specificity for each possible test positivity cut-off. The **area under the ROC curve** informs in a single numerical value about the overall diagnostic accuracy of the index test.

The **intended use** of a medical test can be diagnosis, screening, staging, monitoring, surveillance, prediction or prognosis. The **clinical role** of a test explains its position relative to existing tests in the clinical pathway. A replacement test, for example, replaces an existing test. A triage test is used before an existing test; an add-on test is used after an existing test.

Besides diagnostic accuracy, several other outcomes and statistics may be relevant in the evaluation of medical tests. Medical tests can also be used to classify patients for purposes other than diagnosis, such as staging or prognosis. The STARD list was not explicitly developed for these other outcomes, statistics, and study types, although most STARD items would still apply.

DEVELOPMENT

This STARD list was released in 2015. The 30 items were identified by an international expert group of methodologists, researchers, and editors. The guiding principle in the development of STARD was to select items that, when reported, would help readers to judge the potential for bias in the study, to appraise the applicability of the study findings and the validity of conclusions and recommendations. The list represents an update of the first version, which was published in 2003.

More information can be found on [http://www.equator-network.org/reporting-guidelines/stard](http://www.equator-network.org/reporting-guidelines/stard/).

|  | **Section & Topic** | **No** | **Item** | **Reported on page #** |
| --- | --- | --- | --- | --- |
|  | **TITLE OR ABSTRACT** |  |  | **1** |
|  |  | **1** | Identification as a study of diagnostic accuracy using at least one measure of accuracy  (such as sensitivity, specificity, predictive values, or AUC) | 1 |
|  | **ABSTRACT** |  |  |  |
|  |  | **2** | Structured summary of study design, methods, results, and conclusions  (for specific guidance, see STARD for Abstracts) | 2 |
|  | **INTRODUCTION** |  |  |  |
|  |  | **3** | Scientific and clinical background, including the intended use and clinical role of the index test | 4 |
|  |  | **4** | Study objectives and hypotheses | 5 |
|  | **METHODS** |  |  |  |
|  | *Study design* | **5** | Whether data collection was planned before the index test and reference standard  were performed (prospective study) or after (retrospective study) | 5 |
|  | *Participants* | **6** | Eligibility criteria | 5, table 1 (page 12) |
|  |  | **7** | On what basis potentially eligible participants were identified  (such as symptoms, results from previous tests, inclusion in registry) | 5 |
|  |  | **8** | Where and when potentially eligible participants were identified (setting, location and dates) | 5 |
|  |  | **9** | Whether participants formed a consecutive, random or convenience series | 5 |
|  | *Test methods* | **10a** | Index test, in sufficient detail to allow replication | 6 |
|  |  | **10b** | Reference standard, in sufficient detail to allow replication | 6 |
|  |  | **11** | Rationale for choosing the reference standard (if alternatives exist) | NA |
|  |  | **12a** | Definition of and rationale for test positivity cut-offs or result categories  of the index test, distinguishing pre-specified from exploratory | 5 |
|  |  | **12b** | Definition of and rationale for test positivity cut-offs or result categories  of the reference standard, distinguishing pre-specified from exploratory | NA |
|  |  | **13a** | Whether clinical information and reference standard results were available  to the performers/readers of the index test | 6 |
|  |  | **13b** | Whether clinical information and index test results were available  to the assessors of the reference standard | 6 |
|  | *Analysis* | **14** | Methods for estimating or comparing measures of diagnostic accuracy | 7 |
|  |  | **15** | How indeterminate index test or reference standard results were handled | 7 |
|  |  | **16** | How missing data on the index test and reference standard were handled | 7 |
|  |  | **17** | Any analyses of variability in diagnostic accuracy, distinguishing pre-specified from exploratory | NA |
|  |  | **18** | Intended sample size and how it was determined | 7 |
|  | **RESULTS** |  |  |  |
|  | *Participants* | **19** | Flow of participants, using a diagram | Figure 1 (page 17) |
|  |  | **20** | Baseline demographic and clinical characteristics of participants | Table 2 (page 18) |
|  |  | **21a** | Distribution of severity of disease in those with the target condition | Table 2 (page 18) |
|  |  | **21b** | Distribution of alternative diagnoses in those without the target condition | NA |
|  |  | **22** | Time interval and any clinical interventions between index test and reference standard | 6 |
|  | *Test results* | **23** | Cross tabulation of the index test results (or their distribution)  by the results of the reference standard | Table 3 (page 20) and Supplementary Table 4 |
|  |  | **24** | Estimates of diagnostic accuracy and their precision (such as 95% confidence intervals) | Table 3 (page 20) |
|  |  | **25** | Any adverse events from performing the index test or the reference standard | NA |
|  | **DISCUSSION** |  |  |  |
|  |  | **26** | Study limitations, including sources of potential bias, statistical uncertainty, and generalisability | 11 |
|  |  | **27** | Implications for practice, including the intended use and clinical role of the index test | 12 |
|  | **OTHER INFORMATION** |  |  |  |
|  |  | **28** | Registration number and name of registry | NA |
|  |  | **29** | Where the full study protocol can be accessed | NA |
|  |  | **30** | Sources of funding and other support; role of funders | 12 |

STROBE Statement—Checklist of items that should be included in reports of ***cohort studies***

|  | **Item No** | **Recommendation** | **Page No** |
| --- | --- | --- | --- |
| **Title and abstract** | 1 | (*a*) Indicate the study’s design with a commonly used term in the title or the abstract | 2 |
|  |  | (*b*) Provide in the abstract an informative and balanced summary of what was done and what was found | 2 |
| **Introduction** | | | |
| Background/rationale | 2 | Explain the scientific background and rationale for the investigation being reported | 4 |
| Objectives | 3 | State specific objectives, including any prespecified hypotheses | 5 |
| **Methods** | | | |
| Study design | 4 | Present key elements of study design early in the paper | 5 |
| Setting | 5 | Describe the setting, locations, and relevant dates, including periods of recruitment, exposure, follow-up, and data collection | 5 |
| Participants | 6 | (*a*) Give the eligibility criteria, and the sources and methods of selection of participants. Describe methods of follow-up | 5 and Table 1 |
|  |  | (*b*) For matched studies, give matching criteria and number of exposed and unexposed |  |
| Variables | 7 | Clearly define all outcomes, exposures, predictors, potential confounders, and effect modifiers. Give diagnostic criteria, if applicable | 6 |
| Data sources/ measurement | 8* | For each variable of interest, give sources of data and details of methods of assessment (measurement). Describe comparability of assessment methods if there is more than one group | 7 |
| Bias | 9 | Describe any efforts to address potential sources of bias | 7 |
| Study size | 10 | Explain how the study size was arrived at | 7 |
| Quantitative variables | 11 | Explain how quantitative variables were handled in the analyses. If applicable, describe which groupings were chosen and why | 7 |
| Statistical methods | 12 | (*a*) Describe all statistical methods, including those used to control for confounding | 7 |
|  |  | (*b*) Describe any methods used to examine subgroups and interactions | 7 |
|  |  | (*c*) Explain how missing data were addressed | 7 |
|  |  | (*d*) If applicable, explain how loss to follow-up was addressed | NA |
|  |  | (*e*) Describe any sensitivity analyses | 7 |
| **Results** | | |  |
| Participants | 13* | (a) Report numbers of individuals at each stage of study—eg numbers potentially eligible, examined for eligibility, confirmed eligible, included in the study, completing follow-up, and analysed | Figure 1 |
|  |  | (b) Give reasons for non-participation at each stage | Figure 1 |
|  |  | (c) Consider use of a flow diagram | Figure 1 |
| Descriptive data | 14* | (a) Give characteristics of study participants (eg demographic, clinical, social) and information on exposures and potential confounders | Table 2 |
|  |  | (b) Indicate number of participants with missing data for each variable of interest | Table 2 |
|  |  | (c) Summarise follow-up time (eg, average and total amount) | NA |
| Outcome data | 15* | Report numbers of outcome events or summary measures over time | 9 |

| Main results | 16 | (*a*) Give unadjusted estimates and, if applicable, confounder-adjusted estimates and their precision (eg, 95% confidence interval). Make clear which confounders were adjusted for and why they were included | | | 8 |
| --- | --- | --- | --- | --- | --- |
|  |  | (*b*) Report category boundaries when continuous variables were categorized | | | Table 2 |
|  |  | (*c*) If relevant, consider translating estimates of relative risk into absolute risk for a meaningful time period | | | NA |
|  | Other analyses | 17 | Report other analyses done—eg analyses of subgroups and interactions, and sensitivity analyses | Supplementary table 3 |  |
|  | **Discussion** | | | |  |
|  | Key results | 18 | Summarise key results with reference to study objectives | Table 3 |  |
|  | Limitations | 19 | Discuss limitations of the study, taking into account sources of potential bias or imprecision. Discuss both direction and magnitude of any potential bias | 11 |  |
|  | Interpretation | 20 | Give a cautious overall interpretation of results considering objectives, limitations, multiplicity of analyses, results from similar studies, and other relevant evidence | 12 |  |
|  | Generalisability | 21 | Discuss the generalisability (external validity) of the study results | 12 |  |
|  | **Other information** | | | |  |
|  | Funding | 22 | Give the source of funding and the role of the funders for the present study and, if applicable, for the original study on which the present article is based | 13 |  |

*Give information separately for exposed and unexposed groups.

**Note:** An Explanation and Elaboration article discusses each checklist item and gives methodological background and published examples of transparent reporting. The STROBE checklist is best used in conjunction with this article (freely available on the Web sites of PLoS Medicine at http://www.plosmedicine.org/, Annals of Internal Medicine at http://www.annals.org/, and Epidemiology at http://www.epidem.com/). Information on the STROBE Initiative is available at http://www.strobe-statement.org.
